# Optimizing direct RT-LAMP to detect transmissible SARS-CoV-2 from primary nasopharyngeal swab and saliva patient samples

**DOI:** 10.1101/2020.08.30.20184796

**Authors:** Dawn M. Dudley, Christina M. Newman, Andrea M. Weiler, Mitchell D. Ramuta, Cecilia G. Shortreed, Anna S. Heffron, Molly A. Accola, William M. Rehrauer, Thomas C. Friedrich, David H. O’Connor

## Abstract

SARS-CoV-2 testing is crucial to controlling the spread of this virus, yet shortages of nucleic acid extraction supplies and other key reagents have hindered the response to COVID-19 in the US. Several groups have described loop-mediated isothermal amplification (LAMP) assays for SARS-CoV-2, including testing directly from nasopharyngeal swabs and eliminating the need for reagents in short supply. Here we describe a fluorescence-based RT-LAMP test using direct nasopharyngeal swab samples and show consistent detection in clinically confirmed samples, albeit with approximately 100-fold lower sensitivity than qRT-PCR. We demonstrate that adding lysis buffer directly into the RT-LAMP reaction improves the sensitivity of some samples by approximately 10-fold. Overall, the limit of detection (LOD) of RT-LAMP using direct nasopharyngeal swab or saliva samples without RNA extraction is 1×10^5^-1×10^6^ copies/ml. This LOD is sufficient to detect samples from which infectious virus can be cultured. Therefore, samples that test positive in this assay contain levels of virus that are most likely to perpetuate transmission. Furthermore, purified RNA in this assay achieves a similar LOD to qRT-PCR and we provide a revised method to work directly with saliva as starting material. These results indicate that high-throughput RT-LAMP testing could augment qRT-PCR in SARS-CoV-2 screening programs, especially while the availability of qRT-PCR testing and RNA extraction reagents is constrained.

## Introduction

There are more than 5.8 million reported severe acute respiratory syndrome coronavirus 2 (SARS-CoV-2) infections in the United States as of August 27, 2020 (https://www.cdc.gov/coronavirus/2019-ncov/cases-updates/cases-in-us.html). The actual number of infections is likely far greater since testing remains limited. We now understand that asymptomatic individuals contain similar levels of SARS-CoV-2 in the upper respiratory tract as symptomatic individuals (1-6). Furthermore, 17 out of 24 (71%) presymptomatic patients had positive viral cultures 1 to 6 days before the onset of symptoms (1). Symptom-based screening is not sufficient for controlling SARS-CoV-2 transmission and emphasizes the need for expanded nucleic acid screening of asymptomatic/presymptomatic individuals.

Identifying individuals shedding the most SARS-CoV-2 virus is critical for interrupting transmission. The threshold of virus in a patient sample required to isolate and grow the virus in tissue culture is one indicator of the viral load necessary to transmit the virus. Recent virological assessments of COVID-19 patients suggest that virus isolation from patient samples is dependent on viral load and sample type (1, 7-9). Wölfel et al. (7) demonstrated that successful SARS-CoV-2 isolation was limited to only NP swabs and sputum that had viral loads greater than 1×10^6^ copies of viral RNA (vRNA)/ml (7). This same virus isolation threshold of 1×10^6^ vRNA copies/ml was also found by Van Kampen et al. (10) when analyzing 129 severe COVID-19 patient samples and Quicke et al. (11) in a longitudinal surveillance study of 454 skilled nursing facility staff members. During a SARS-CoV-2 outbreak in a Washington nursing home, virus could be isolated only from NP swabs with RT-PCR cycle threshold (Ct) values of less than 30, with few exceptions, collected from patients presenting as asymptomatic, presymptomatic, with typical and atypical symptoms (1). A similar virus isolation threshold (Ct value 33-34; approximately 1×10^5^ RNA copies/ml) was observed in SARS-CoV-2 patients by La Scola et al. (2020) when using a qRT-PCR assay targeting E gene (12). In a recent non-human primate study, virus was successfully isolated from the upper respiratory tract of rhesus macaques inoculated with 2.6×10^6^ TCID50 of SARS-CoV-2 via a combination of intratracheal, intranasal, ocular, and oral routes (9). Positive viral cultures were limited to NP and oral swabs with greater than 1×10^6^ and 1×10^5^ copies/ml respectively (9). These data suggest that diagnostic tests with a sensitivity around 1×10^6^ copies/ml are sufficient for capturing culture-positive cases with the greatest transmission risk.

Conventional SARS-CoV-2 testing relies on RT-PCR amplification of virus-specific nucleic acids extracted from nasopharyngeal (NP) swabs. However, shortages of nucleic acid extraction and RT-PCR reagents as well as RT-PCR instrumentation remain a problem (13). Alternative nucleic acid extraction methods and “direct” testing that does not require nucleic acid extraction are important to expand testing while reducing time and cost. Indeed, the SalivaDirect method recently approved under an FDA EUA, utilizes saliva without RNA extraction into a RT-PCR assay, eliminating at least part of the process experiencing shortages (14).

Loop-mediated isothermal amplification (LAMP) has been used as a tool for point-of-need diagnostic testing for several pathogens, including SARS-CoV-2 (15-24). LAMP assays are an alternative method for rapidly detecting the presence of specific nucleic acids in samples, with colorimetric or fluorescent visualization of results. LAMP assays are inexpensive, high-throughput, do not necessarily require nucleic acid purification, and give rapid results. These previously published manuscripts demonstrate proof-of-principle for SARS-CoV-2 testing by RT-LAMP using either contrived samples with free nucleic acid or primary samples with RNA isolation first.

In this study, we focused on characterizing and optimizing direct RT-LAMP without RNA isolation and with primary NP swab samples with known SARS-CoV-2 status. We demonstrate the limit of detection (LOD) of direct swab RT-LAMP in primary swab samples as well as modifications of the technique that help improve sensitivity, but don’t rely on the same materials required for traditional qRT-PCR methods. We characterized the use of Lucigen QuickExtract (QE) lysis buffer, guanidine hydrochloride addition, an alternative RNA isolation method, and several primer sets and combinations targeting different gene regions. Lastly, we used our optimized approach to test the transition to direct RT-LAMP with saliva in a point-of-need testing approach. Each of these modifications are useful additions to the SARS-CoV-2 testing repertoire and have unique benefits for testing in multiple laboratory settings. Furthermore, we suggest that the limit of sensitivity achieved with any of these methods is sufficient to detect levels of virus that can be cultured out of samples and therefore represents levels where transmission is most likely and self-quarantine most important.

## Materials and Methods

### Sample collection

Residual NP swab and saliva samples were provided by University of Wisconsin-Madison Hospitals and Clinics and the Wisconsin State Laboratory of Hygiene under biosafety protocol B00000117 (IRB 2016-0605) and their use was not considered human subjects research by the University of Wisconsin-Madison School of Medicine and Public Health’s Institutional Review Board. Samples were collected into a variety of transport media including universal transport media (UTM), viral transport media (VTM), and phosphate buffered saline (PBS), stored at 4°C for up to 7 days, and transported to the laboratory at room temperature. Upon arrival at the laboratory, samples were stored at either 4°C (for immediate same-day use) or −80°C until use in RT-PCR or RT-LAMP assays. Residual saliva samples contained no media/buffer and were stored for 2-4 weeks at 4°C before arriving in the lab and were tested within 2 days of arrival.

### qRT-PCR

Viral load analysis was performed after samples arrived in our laboratory. RNA was isolated using the Viral Total Nucleic Acid kit for the Maxwell RSC instrument (Promega, Madison, WI) following the manufacturer’s instructions. Saliva samples were diluted 1:1 in water prior to extraction. Viral load quantification was performed using a qRT-PCR assay developed by the CDC to detect SARS-CoV-2 (specifically the N1 assay) and commercially available from IDT (Coralville, IA). The assay was run on a LightCycler 96 or LC480 instrument (Roche, Indianapolis, IN) using the Taqman Fast Virus 1-step Master Mix enzyme (Thermo Fisher, Waltham, MA). The LOD of this assay is estimated to be 200 genome equivalents/ml saliva or swab fluid. To determine the viral load, samples were interpolated onto a standard curve consisting of serial 10-fold dilutions of *in vitro* transcribed SARS-CoV-2 N gene RNA kindly provided by Nathan Grubaugh (Yale University).

### RT-LAMP

The experiments we describe here were modified from the SARS-CoV-2 RT-LAMP assay developed by Zhang et al. (16). For most assays we used fluorescent-based detection with Warmstart LAMP reagents and the included fluorescent dye (New England Biolabs, NEB). We tested primer sets developed in previous studies targeting several SARS-CoV-2 genes as shown in S1 Table (16, 17, 25-29). Of note, the Color-Orf1a primers and Lamb-Orf1a primers are identical, but were used at different concentrations per the protocols developed by each lab. The final 1X primer concentrations are listed in the table. For each reaction, a 10X stock of all 6 primers were combined with Warmstart mastermix and water in 25μl reactions following the manufacturer’s recommendations. Unless otherwise stated, 1μl RNA transcript of the SARS-CoV-2 N-gene obtained by Dr. Nathan Grubaugh, 1μl of synthetic SARS-CoV-2 RNA transcript (Twist Biosciences; RNA control 2), 1μl of gamma-irradiated SARS-CoV-2 (BEI; NR-52287; isolate USA-WA1/2020), or 1μl primary NP swab sample were tested in each RT-LAMP reaction. Unless otherwise stated, all serial dilutions were performed in water. For reactions testing guanidine hydrochloride addition to the RT-LAMP mastermix, a final concentration of 40mM stock was used in the mastermix. Except where otherwise specified, samples were run on a Roche Lightcycler 96 instrument (Roche Diagnostics) using an 80-cycle program with the SYBR Green channel at 65°C (495-497 nm absorption; 517-520 nm emission) and data collection every 30 seconds. For experiments determining the appropriate volume of direct swab sample addition for highest RT-LAMP efficiency, a 60-cycle program with data collection every 20 seconds was used. For colorimetric RT-LAMP with saliva, saliva was incubated at 65°C for 30 minutes followed by 98°C for 3 minutes in a heat block. Saliva was then diluted 1:1 in PBS and 3μl was added into a 20μl RT-LAMP mastermix containing the Color-N primer set and colorimetric mastermix (NEB) and incubated at 65°C for 30 minutes.

### Sample lysis

A subset of samples was treated with LucigenQE RNA Extraction Solution (Lucigen, Middleton, WI) in a 1:1 ratio as described in Ladha et al. (30). Briefly, NP swab eluate was combined with an equivalent volume of LucigenQE and briefly vortexed. Samples were then incubated for 5 minutes at 95°C, cooled on ice, and maintained until addition to the RT-LAMP reaction.

### Statistical analysis

To assess improvement in quantification cycle (Cq) values using sample lysis with LucigenQE or RNA isolation, mean Cq values were calculated for each sample. Mean Cq values were not normally distributed for either dataset so we used a nonparametric equivalent to a paired t-test, the Wilcoxon signed rank test with continuity correction, for each set of paired samples.

To examine whether sample vRNA load and/or treatment were significantly associated with a positive RT-LAMP result, we used logistic regression with RT-LAMP result as the dependent variable. For our analysis, an equivocal result in which one replicate was positive while the other was negative, was conservatively treated as negative. We coded RT-LAMP results for each sample tested by each method as a dichotomous outcome with positive samples coded as “1” and negative or equivocal samples coded as “0”. For explanatory variables, we chose qRT-PCR vRNA load, with samples greater than 10^6^ copies/ml coded as “1” and samples less than 10^6^ copies/ml coded as “0”, and group, designated as either 5μl RNA, 1μl lysed, or 1μl direct addition.

All statistical analyses were performed in RStudio (v. 1.2.1335) using R (v. 3.6.0) (31).

## Results

### Limit of detection with RNA transcript and Gene-N-A primers

To determine a limit of detection for the RT-LAMP assay, serial 10-fold dilutions of RNA transcript containing the N-gene were tested in RT-LAMP reactions with Gene-N-A primers in duplicate in 3 independent assays. RNA transcripts were diluted in RNase-free water. Consistent detection of RNA was achieved when 1×10^6^ copies or greater of RNA/ml was added into the reactions (1×10 copies/μl per reaction) (Fig 1A). To obtain a more precise LOD, transcript was diluted 1:2 starting at 5×10^6^ copies/ml down to 7.8×10^4^ copies/ml and each concentration was run in 10 replicates. Nine of ten replicates at 6.25×10^5^ copies/ml were positive, while 6/10 were positive at 3.12×10^5^ copies/ml and 5/10 were positive at 1.56×10^5^ copies/ml (Fig 1A). Thus, we can consistently detect 625 copies of input into the reaction, but can detect down to 156 copies of input in half of the reactions. Zero reactions were detected as positive at 1×10^5^ copies/ml (100 copies/reaction) or below.

**Fig 1:**
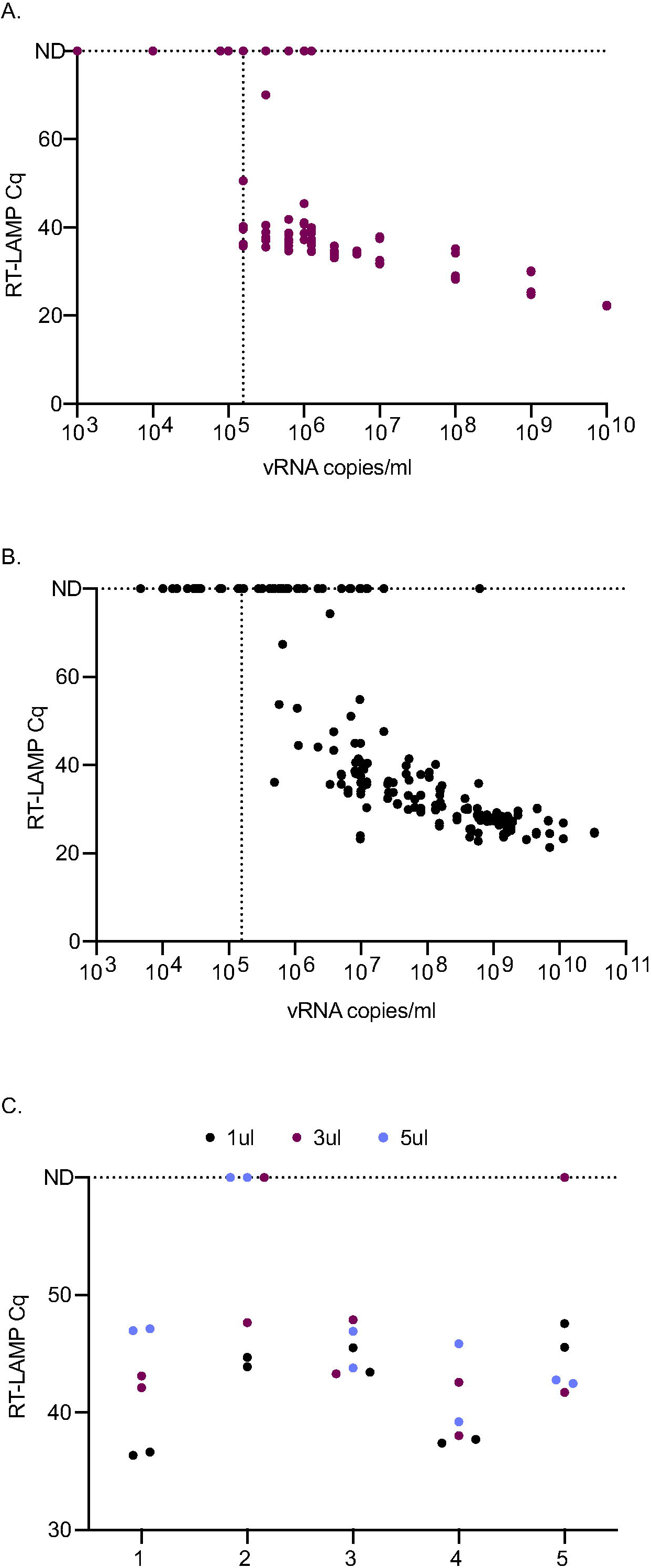
Detection of SARS-CoV-2 by RT-LAMP from transcript or primary NP swab samples. A. The quantification cycle (Cq) relative to each transcript copy number is plotted. Samples that were not detectable were plotted on the line labeled ND for all graphs at Cq of 60 or 80, the total number of cycles run in our assays. The vertical line is set at the lowest dilution where positive samples were detected using the transcript input for all graphs (156,300 vRNA copies/ml). Each replicate is plotted on all graphs. B. Detection of 106 SARS-CoV-2 positive primary NP swab samples relative to their in-house viral load value. C. RT-LAMP Cq of five SARS-CoV-2 positive primary NP swab samples with different swab input volumes.

### Limit of detection with primary nasopharyngeal swab samples

Leftover NP swab samples from 106 patients with known clinical diagnosis of SARS-CoV-2 were tested directly by RT-LAMP in duplicate. Additionally, RNA was isolated from these samples and tested by qRT-PCR with a transcript standard for quantitation. A total of 63/106 (59%) samples tested positive by RT-LAMP and 106/106 by qRT-PCR (S2 Table). Another 13 samples were equivocal by RT-LAMP, with one of two replicates positive. As shown in Fig 1B, the LOD of primary samples was similar to that seen with RNA transcript. 63/77 (82%) samples with viral RNA copy numbers greater than 1×10^6^ copies/ml were detected by RT-LAMP, whereas 0/28 samples with concentrations <1×10^6^ were detected positive and 3/28 were equivocal. All 31 samples that tested negative for SARS-CoV-2 by clinical laboratories also tested negative by RT-LAMP (data not shown).

### RT-LAMP is inhibited by adding larger volumes of primary sample

To determine whether adding larger volumes of primary NP swab samples could improve the sensitivity of the assay, 1, 3, and 5μl of swab samples from 5 primary SARS-CoV-2-positive samples were tested side-by-side. All replicates were detected as positive when 1μl was added directly into the RT-LAMP reaction (Fig 1C). However, one of the two replicates from two samples tested negative when 3μl of sample was added and both replicates from one sample tested negative when 5μl of sample was added into the RT-LAMP reaction. Furthermore, Cq thresholds were higher with addition of higher volumes of sample. Therefore, we chose to use 1μl of straight swab sample in subsequent experiments.

### Lysis buffer improves the sensitivity of RT-LAMP

To determine whether treatment with lysis buffer improves the sensitivity of RT-LAMP to detect SARS-CoV-2 in clinical samples without the use of traditional nucleic acid isolation methods, we treated 72 clinical samples with a range of SARS-CoV-2 vRNA loads in a 1:1 ratio with LucigenQE as described by Ladha et al. (30). We then compared the fluorescent RT-LAMP Cq values between 1μl of lysed sample and 1μl of the same samples added directly. Addition of 1μl of NP swab eluate directly into the RT-LAMP reaction resulted in positive detection in both replicates of 46/72 (64%) known SARS-CoV-2-positive samples and in 1 of 2 replicates in 6 additional samples (Fig 2A, S2 Table). Treatment of the same 72 samples with LucigenQE resulted in detection of both replicates for 56/72 (78%) samples, an additional 10 samples that were undetectable before. An additional 4 samples that were negative when tested directly were equivocal when treated with LucigenQE. Negative samples with either method occurred in samples with viral loads ranging from 1.0×10^4^ −6.19×10^8^ vRNA copies/ml. However, while 0/19 samples with viral loads below 1×10^6^ were positive with 1μl straight, 4 of 19 samples were detectable after lysis in both replicates and 4 additional samples were detected in 1 of 2 replicates. Mean Cq for LucigenQE-treated samples were significantly lower (Cq = 38.62) than those for directly added samples (Cq = 49.08) (Wilcoxon signed rank test, V = 1830, p = 1.67E” ^11^), suggesting that lysis treatment improves the efficiency of amplification in the LAMP reaction (Fig 2A). We also examined whether sample vRNA load and/or treatment were significantly associated with RT-LAMP detection results. We found that direct addition of 1μl of untreated NP swab was associated with a decreased odds of detecting a positive RT-LAMP result (OR = 0.20, 95%CI = 0.04-0.65, p = 0.015) while the most important factor associated with a positive result was a sample vRNA load of greater than 10^6^ copies/ml (OR = 88.5, 95%CI = 25.74-434.87, p = 1.88E^-10^).

**Fig 2:**
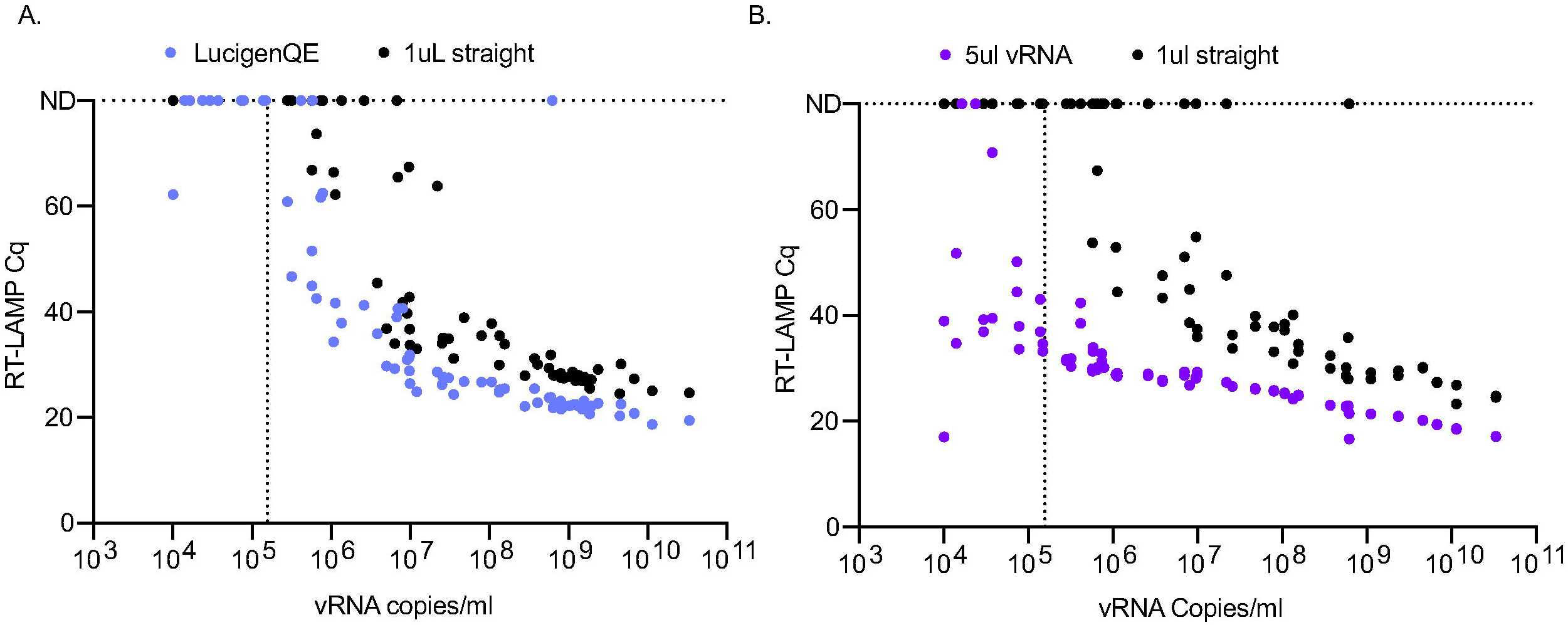
Comparison of the detection of SARS-CoV-2 positive primary NP swab samples after using direct sample addition to either LucigenQE treatment or isolated vRNA. A. Comparison of Cq values between samples treated with or without LucigenQE and run by RT-LAMP with 1μl of sample. B. Comparison of Cq values between RT-LAMP assays run with 1μl straight swab or 5μl of isolated and purified vRNA.

### RNA isolation improves the limit of detection of RT-LAMP to levels similar to qRT-PCR

One of the primary reasons the direct RT-LAMP assay is less sensitive than the diagnostic qRT-PCR assay is because the qRT-PCR assay uses 5μl of concentrated and purified RNA as input. To determine whether the RT-LAMP assay would perform to a similar level of detection if the same input was used, 5μl of purified RNA was used in RT-LAMP assays from a subset (n=44) of COVID-19-positive NP swab samples. The viral loads ranged from 1.01×10^4^ to 1.14×10^10^. 19 of 44 had concentrations of virus below the RT-LAMP LOD of 1×10^6^ vRNA copies/ml. Of the 44 samples tested with 1μl of direct swab, 18 tested positive (43%) and 7 were equivocal between replicates (Fig 2B and S2 Table). When 5μl of purified RNA was used in the reactions instead, 42 of 44 samples (95%) tested positive (Fig 2B, S2 Table). Furthermore, samples with 100-fold lower vRNA copies/ml were detected after RNA isolation by RT-LAMP. Adding purified RNA brings the possible LOD of detection down to 50 copies of input of a 10 copy/μl sample (1×10^4^ copies/ml), which was detected in 4 of 6 primary samples tested within this viral load range. Overall, the addition of 1μl NP swab direct was associated with reduced odds of a positive RT-LAMP result (OR = 0.0054, 95%CI = 0.00023-0.041, p = 2.56E^-05^). Similar to the results for lysis buffer treatment, samples with qRT-PCR vRNA loads greater than 1×10^6^ copies/ml had significantly increased odds of a positive RT-LAMP result (OR = 49.35, 95%CI = 8.22-966.29, p = 0.00044). Lastly, the Cq values were lower for all detected samples when 5μl of RNA was added (mean Cq = 31.42) instead of 1μl straight sample (mean Cq = 58.72), indicating faster and more robust detection with concentrated and purified RNA (Wilcoxon signed rank test, V = 903, p = 1.71E^-08^) (Fig 2B).

### Alternative primers improve efficiency

Multiple SARS-CoV-2 RT-LAMP primer sets have been published or included in manuscripts on pre-print servers since we began our experiments. We compared the efficiency of several alternative primer sets either alone or in combination to the Gene-N-A primers used in our initial studies (S1 Table). Two primary NP swab samples with high concentrations of SARS-CoV-2 (NP1:1.09×10^12^ copies/ml, NP2:4.28×10^10^ copies/ml) as well as gamma-irradiated SARS-CoV-2 (BEI) were used to screen different primer combinations using the same fluorescent RT-LAMP conditions. First, a set of primers previously established (17, 27) and used to obtain an FDA EUA by Color Genomics was tested with each primer alone and in different combinations (Fig 3A). Several primers and primer combinations resulted in a lower Cq value across the board than the Gene-N-A gene primers suggesting improved reaction efficiency. The primer combinations including Color-N/Color-E and Color-N/Color-ORF1a yielded the lowest Cq values in all samples.

**Fig 3:**
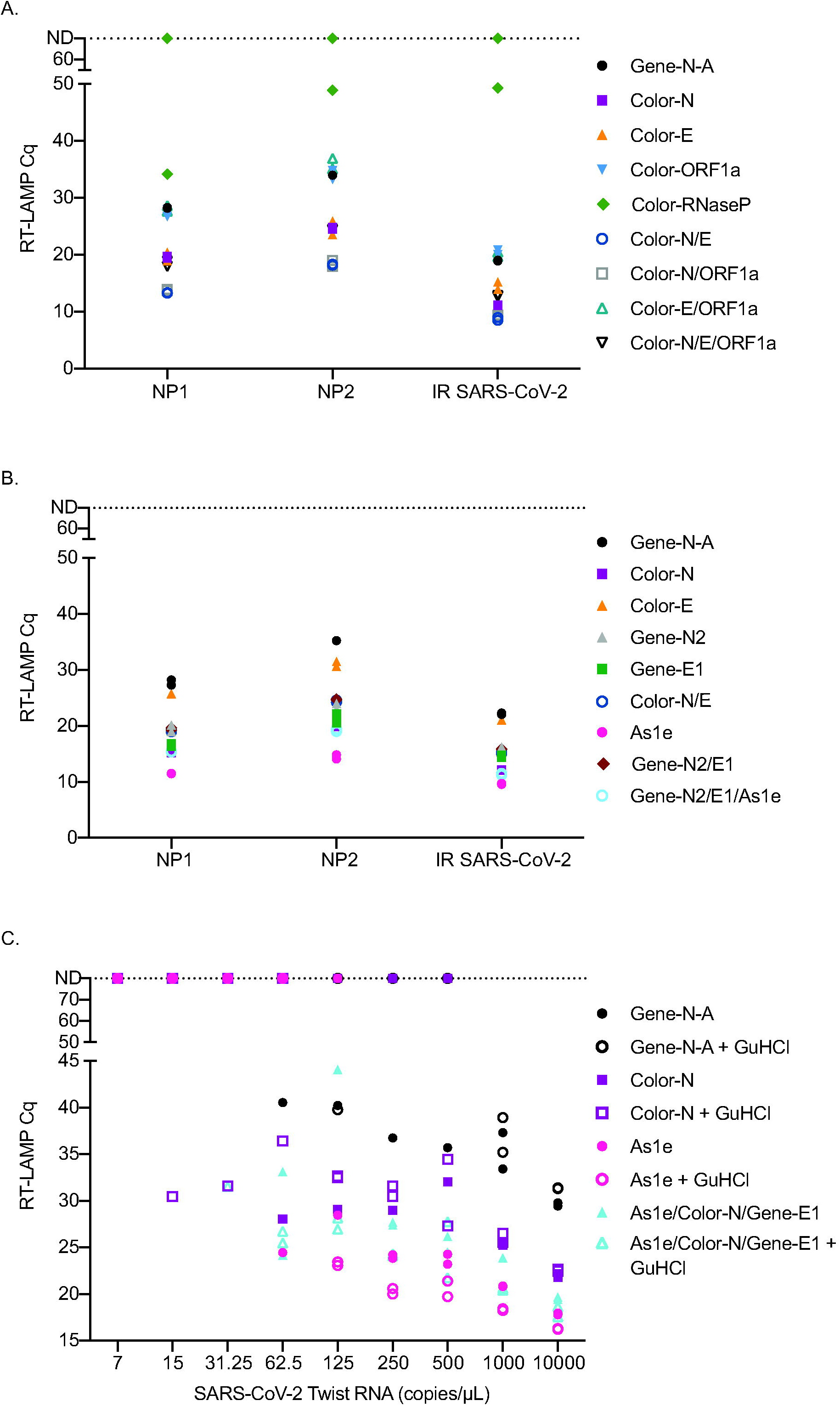
Comparison of RT-LAMP Cq value on primary NP swab samples, irradiated SARS-CoV-2 or SARS-CoV-2 TWIST RNA amplified with different primer sets. Reactions were run in duplicate and both replicates are shown on the graphs. Samples that were not detectable are plotted on the ND line at Cq 80. A. Comparison of Cq values using Color Genomics primers to the Gene-N-A primers on two primary NP swab samples and irradiated SARS-CoV-2. B. Comparison of Zhang et al.(25) primers to a subset of Color Genomics primers and Gene-N-A primers on two primary samples and irradiated SARS-CoV-2. C. Comparison of the Cq values obtained when using the best primers and combinations of primers across different dilutions of SARS-CoV-2 TWIST RNA with and without GuHCl.

Next, we compared the Gene-N-A, Color-N, Color-E, and Color-N/E combination to second generation primers described in Zhang et al. (25) (Gene-N2, Gene-E1 and As1e). We found that As1e, originally published by Rabe et al. (26) yielded the lowest Cq value followed closely by a combination of As1e with the two primers targeting the Gene-N2 and Gene-E1 genes designed by Zhang et al. (Fig 3B). The Color-N primer yielded similar Cq values to the triple combination primer set from Zhang et al. We also tested the Color-N, Color-E1, and Gene-N-A gene primer sets against additional published primers that target ORF1a (Lamb, Yu, El-Tholoth, and Zhang primers) either alone or in combination and compared them to the Gene-N-A gene primer set (S1 Table). The Color-N primer produced the lowest Cq value in this set (S1 Fig).

We then tested As1e, Color-N, As1e/Color-N/Gene-E1 primer set with and without guanidine hydrochloride, as recommended by Zhang et al., with Twist RNA SARS-CoV-2 template. Under these conditions, guanidine hydrochloride improved detection with most primer sets with the exception of the Gene-N-A primer set (Fig 3C). Using the Twist RNA, the primer set that detected samples at the lowest dilutions was the Color-N primer or combination of As1e/Color-N/Gene-E1 with guanidine hydrochloride, though only in one of two replicates. The As1e primer with and without GuHCl as well as the combination As1e/Color-N/Gene-E1 with GuHCl often yielded the lowest Cq value.

Lastly, to determine which primer set or combination worked best with primary NP swab samples, fourteen samples with viral loads ranging from 7.33×10^7^-1.52×10^11^ vRNA copies/ml were tested using 1μl of straight swab sample with the most promising primer combinations with and without GuHCl. Both the Color-N primer set alone and the As1e/Color-N/Gene-E1 combination yielded the lowest Cq value across the different levels of virus (Fig 4A). Guanidine hydrochloride did not improve detection in primary samples as seen with the Twist RNA. Twelve additional primary NP swab samples with high Ct values ranging from 25 to 35 were tested with the Color-N and As1e/Color-N/Gene-E1 combination. Detection of these samples was very similar between the two primer conditions (Fig 4B).

**Fig 4:**
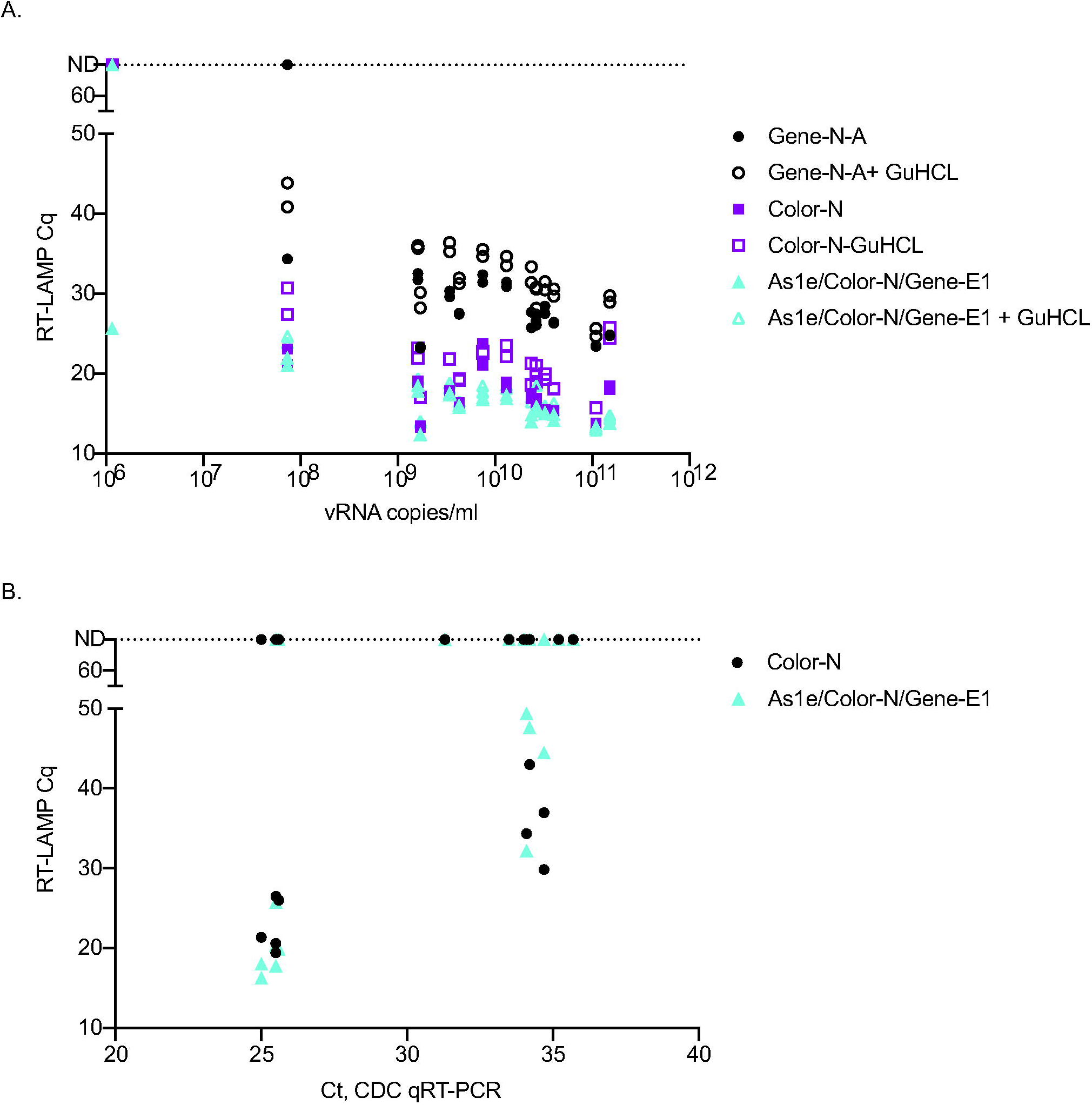
Comparison of the best-performing primers and combinations on additional primary NP swab samples with varying levels of virus with and without GuHCl. A. Comparison of three primer sets or combinations either with or without GuHCl on 14 primary NP swab samples with high viral load copy numbers. B. Comparison of the Color-N primer set to As1e/Color-N/Gene-E1 on primary NP swab samples with lower levels of virus. These samples were not run by our in-house viral load test and therefore Ct value obtained from the hospital is reported.

### Transition to saliva-based direct RT-LAMP

To best accommodate point-of-need testing, saliva-based tests using colorimetric read-out minimizes expensive equipment and eases sample collection. We modified our direct RT-LAMP approach to accommodate detection of SARS-CoV-2 from saliva while also incorporating heat inactivation of the virus for safety in the field (32). Leftover paired NP swab and saliva samples were tested by their respective RT-LAMP protocols as well as by qRT-PCR (Fig 5). Detection of SARS-CoV-2 by RT-LAMP was dependent on the viral load of the sample rather than the starting sample material. Overall, samples with viral loads above 5×10^5^ copies/ml were detected as positive by RT-LAMP from either NP swab or saliva. Viral loads from NP swab samples were often higher than those in saliva in this sample set. This is likely due to the saliva samples being stored for 2-4 weeks at 4°C before use in this assay while NP swabs were stored for 1-2 weeks at 4°C and then were frozen before use. This data should not be used to assess differences in viral load observed between saliva and NP swabs because of the differential storage of these leftover samples. This data does provide proof-of-concept that saliva can be added directly to colorimetric RT-LAMP reactions and yields a LOD similar to NP swab samples.

**Fig 5:**
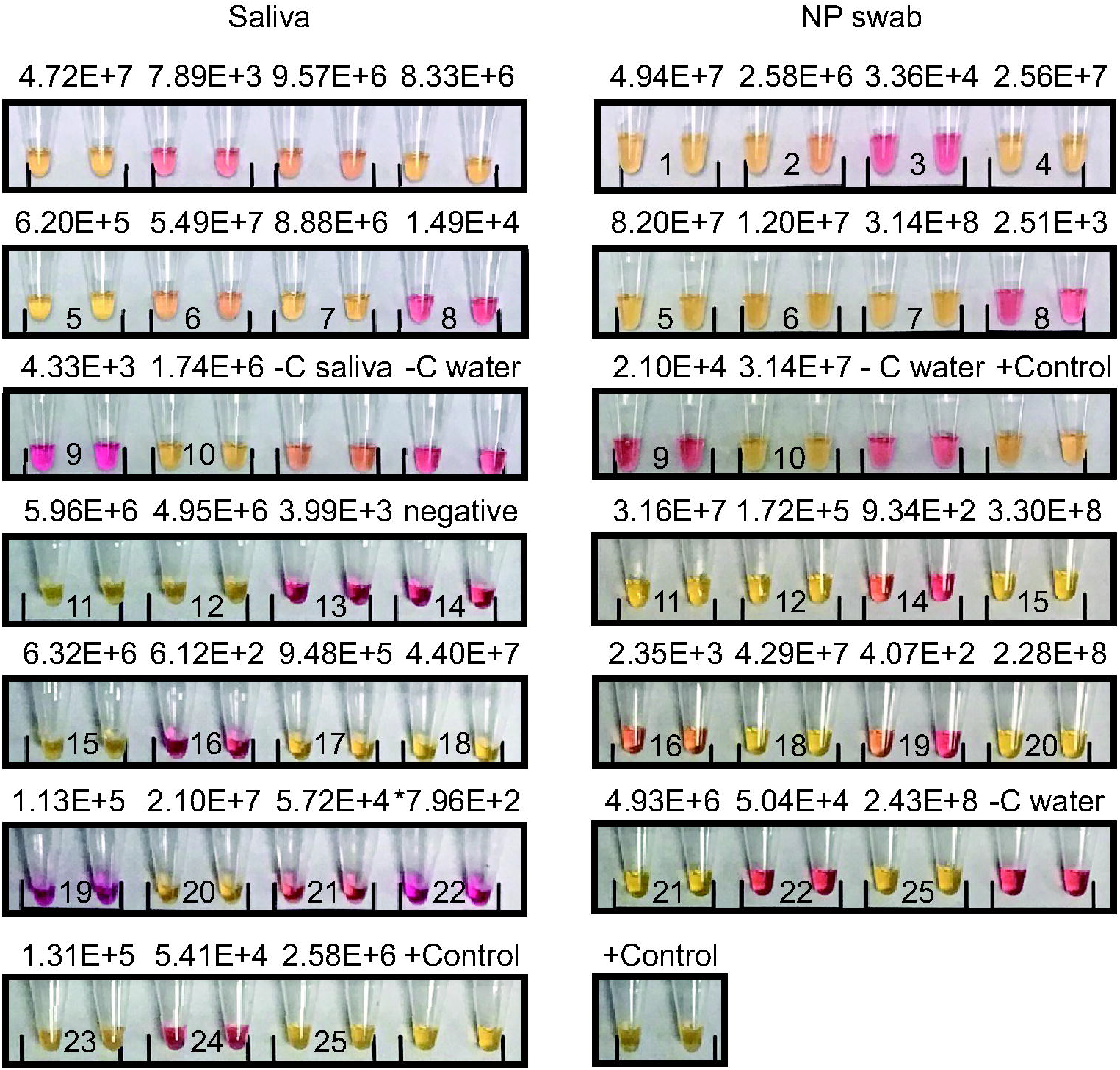
Colorimetric RT-LAMP from SARS-CoV-2 positive paired NP swab and saliva samples. Positive samples are yellow and negative samples are red. qRT-PCR-based viral loads are presented above each sample pair. Paired numbers are from the same patient. Some saliva samples did not have paired NP swab samples. Irradiated SARS-CoV-2 was used as a positive control.

## Discussion

Frequent, widespread testing is considered the best mitigation strategy to control SARS-CoV-2 before a vaccine or effective therapeutic is available. Traditional qRT-PCR is relatively sensitive, but it is time consuming and reliant on very specific reagents that are in short supply in the ongoing pandemic. RT-LAMP has become a promising alternative to qRT-PCR, but the true sensitivity of this assay has been poorly characterized from primary NP swab samples when added directly into the reaction. Many published studies establish a LOD based on free RNA transcript or isolated RNA from cultured virus of around 100 copies/rxn. These LODs apply to RT-LAMP only when purified RNA is used as input. They do not apply to direct RT-LAMP methods containing whole virions in primary samples also containing host enzymes and other host components. In this study we established that 625 copies/reaction was necessary in order to detect RNA transcripts consistently in 9/10 reactions in our assay. In primary samples we rarely detected virus in samples with a vRNA load of less than 5×10^5^-1×10^6^ vRNA copies/ml, establishing this threshold as a conservative LOD. While not as sensitive as methods with RNA extraction first, this LOD range is sufficient for SARS-CoV-2 surveillance to detect virus in individuals with the minimal amount of virus necessary to isolate the virus and therefore most likely to transmit the virus.

The LOD of 1×10^6^ copies/ml is likely due to inefficiencies associated with virus lysis at 65°C during the LAMP reaction and possible degradation of liberated RNA by enzymes, including RNases, present in primary samples. Indeed, adding more sample, including more host enzymes and media with potential inhibitors, reduced detection. On the other hand, lysis with LucigenQE was compatible with RT-LAMP and improved our sensitivity to detect SARS-CoV-2 in primary samples with vRNA loads less than 10^6^ copies/ml. When compared with direct addition, we were able to increase our detection of true positives for both replicates from 61% to 78% after lysis treatment. Guanidine hydrochloride has also been shown to improve sensitivity in other studies and while our results showed better detection with synthetic Twist RNA, bringing the LOD down to 62.5 copies/μl, the same improvement was not observed in primary samples with intact virus. The largest increase in sensitivity occurred with RNA isolation prior to RT-LAMP using an alternative RNA isolation method to those approved for the CDC qRT-PCR assay. With RNA isolation we detected 95% of the samples detected by qRT-PCR, including those with the lowest viral loads.

There are now many primers available targeting different regions of SARS-CoV-2. In this work we tested several sets to iteratively choose which primer set worked best with primary NP swab samples. We found that several primer sets performed more efficiently than the Gene-N-A primers we used in most of our experiments. The two best performing primer sets that were nearly indistinguishable in performance with both high and low viral load primary samples were a combination of As1e/Color-N/Gene E1 and the Color-N primer set alone.

For most of this study we chose to use fluorescent RT-LAMP for detection rather than colorimetric detection. Using fluorescence enabled analysis of the differences in the Cq values providing a quantitative evaluation of how each condition changed the efficiency of the assay. However, when considering how to deploy RT-LAMP in the field, our group has developed a mobile RT-LAMP workflow that uses saliva and colorimetric readouts for low cost and portability. Additional work needs to be done to determine whether the benefits of fluorescent detection can be inexpensively migrated to decentralized point-of-need testing.

Many studies are comparing RT-LAMP to RT-PCR results presented as Ct value, rather than vRNA copies/ml. We chose to focus on comparing methods and determining LODs based on a standard qRT-PCR assay performed in our laboratory with a quantitative standard, rather than Ct values generated by the varying sources of our primary samples. The hospital laboratories have transitioned between different methods as reagents were available and as new assays became available, which means that the Ct values we obtained for each of our samples was generated by different assays targeting different regions of the SARS-CoV-2 genome. By comparing all our results to the vRNA copies/ml that we generated in our lab using a consistent primer set and protocol (CDC qRT-PCR assay) that targeted the same gene (and primers) as our SARS-CoV-2 RT-LAMP assay, we were able to ensure our comparisons were consistent across all samples.

Overall, we have shown that direct RT-LAMP using fluorescent detection can detect SARS-CoV-2 in primary NP swab samples with viral loads greater than 1×10^6^ vRNA copies/ml. We were able to improve this slightly with the quick and low-cost addition of Lucigen lysis buffer to the reaction. We also saw improvement in efficiency with several alternative primer sets. While direct RT-LAMP is not as sensitive as qRT-PCR, the gold standard for diagnosis, it is sufficient to detect the levels of virus that are necessary to culture virus from a sample as described previously. This means that this assay detects people who are likely to transmit the virus for a significant reduction in cost, time, and reagents relative to qRT-PCR. One proposed way to utilize this test could be to screen large numbers of individuals who are then directed toward diagnostic testing by qRT-PCR if they test positive by this test, significantly reducing the burden on diagnostic labs and their resources. Lastly, we confirmed that saliva was also compatible with direct RT-LAMP assays when heated first to inactivate the virus and nucleases. Overall, direct RT-LAMP is an important addition to the repertoire of currently available tests to identify samples containing SARS-CoV-2 nucleic acids.

## Data Availability

All data supporting the results are presented either in the main paper or as part of the supplemental online material.

## Acknowledgements

We thank Nathan Tanner for discussions about primers and optimizations of the RT-LAMP assay. The following reagent was deposited by the Centers for Disease Control and Prevention and obtained through BEI resources, NIAID, NIH: SARS-Related Coronavirus 2, Isolate USA-WA1/2020, Gamma-Irradiated, NR-52287.

## Supporting information captions

**S1 Figure. Comparison of Gene-N-A, Color-N, and Color-E primers to several ORF1a - targeting primer sets and combinations with two primary samples and irradiated SARS-CoV-2**. Samples that were not detectable were plotted on the ND line set at Cq 80, the highest cycle number in our assay.

**S1 Table. Primer sequences and final 1X concentrations used in the RT-LAMP reactions**.

**S2 Table. qRT-PCR N-gene viral loads and RT-LAMP Cq values from 106 primary NP swab samples run in duplicate with 1ul of swab sample or a subset of NP swab samples run in duplicate with either 1ul of primary samples treated with Lucigen QuickExtract or 5ul of extracted vRNA**.

